# Miniaturization of Epileptic Abnormal Electrocorticogram Detector Using 3D Convolutional Neural Network

**DOI:** 10.1101/2025.02.03.25321462

**Authors:** Moemi Yamaji, Shinjiro Yamamasu, Yuto Hirano, Yuki Hayashida

## Abstract

Epilepsy is a neurological disorder characterized by sudden and recurrent seizures caused by abnormal electrical activity in the brain. Responsive Neurostimulation (RNS) offers a promising treatment option for patients with drug-resistant epilepsy. Responsive Neurostimulation (RNS) is an implantable device that employs a closed-loop system. It continuously monitors brain activity through electro-corticogram (ECoG) recordings. When the system detects seizure activity, it delivers direct electrical stimulation to the brain to suppress the seizure. Seizure detection algorithms require patient-specific optimization, leading to increased interest in deep learning approaches in recent years. While deeper network architectures generally improve detection accuracy, their implementation in implantable devices is constrained by limited hardware resources and the restricted number of electrode channels available for ECoG monitoring. To ensure the practical feasibility of RNS, it is crucial to systematically minimize both the computational costs of patient-specific deep learning models and the number of connected ECoG electrodes. This study systematically reduced the number of electrode channels and computational costs in seizure detection models by analyzing the spatiotemporal kernels learned by the first convolutional layer of a 3D convolutional neural network (3D CNN) trained on 3D ECoG data. This approach capitalizes on the network’s ability to learn spatial relationships between grid electrodes and the temporal dynamics of ECoG signals. The performance comparison between the downsized seizure detection CNN model and the original CNN model revealed that, for at least some patients, it is possible to maintain inference performance while reducing the model size.

## I. Introduction

Epilepsy is a neurological disorder characterized by recurrent and unpredictable disruptions in normal brain activity [1]. It is a common condition, affecting approximately 1% of the global population, or about 65 million people worldwide [2]. The primary treatment for epilepsy is pharmacotherapy using antiepileptic drugs to suppress seizures. However, approximately 30% of epilepsy patients are resistant to medication [3]. This drug-resistant epilepsy can lead to psychological distress and severe complications, potentially life-threatening. Consequently, the quality of life (QOL) for these patients is significantly diminished.

Among patients with drug-resistant epilepsy, only a small percentage (approximately 7–8%) are eligible for surgical resection. If the epileptogenic zone (EZ), the area of the brain responsible for seizures, can be accurately localized, surgical resection of the EZ may offer the possibility of seizure freedom. Resection surgery has the potential to cause irreversible damage to neurological functions, necessitating careful consideration [4]. Neurostimulation therapy, an alternative approach for drug-resistant epilepsy, involves delivering electrical stimulation to the brain regions where seizures originate [5]. Responsive Neurostimulation (RNS) [6] is an implantable, closed-loop neuromodulation system. It continuously monitors electro-corticogram (ECoG) activity in real-time and automatically delivers mild electrical stimulation to the affected brain region upon detecting the onset of a seizure, aiming to suppress the epileptic episode [7].

Due to the computational resource limitations of implanted devices, traditional seizure detection algorithms have relied on simple methods like feature thresholding [8]. However, feature thresholding requires extensive manual feature selection and rule design, as the optimal thresholds vary depending on the type of seizure and the individual patient’s brainwave patterns. This process demands significant time and expertise [9], making it challenging to develop an optimized system tailored to each patient. In recent years, deep learning (DL) technology has been increasingly adopted for the development of patient-specific seizure detection algorithms [10][11]. DL models excel at analyzing complex data and have the capability to address challenges such as handling imbalanced datasets and quantifying the uncertainty associated with seizure predictions. Therefore, DL models hold promise for the development of practical tools for epilepsy seizure detection and prediction in the near future [12]. Among the DL architectures used for seizure detection, 2D-CNN is the most widely employed [12]. CNNs are particularly adept at capturing local spatial dependencies, making them well-suited for extracting important patterns from ECoG signals. 3D-CNNs excel at capturing intricate spatiotemporal patterns within ECoG data, incorporating both the spatial relationships between electrodes and the temporal dynamics of brain activity. This potential for enhanced performance comes at the cost of increased computational demands, requiring significant hardware resources for training. Additionally, while deeper network layers generally enhance detection accuracy, they also lead to higher computational costs. Consequently, integrating DL-based seizure detection algorithms into clinical-grade devices, which require real-time processing and high energy efficiency, remains a significant challenge. Liu and Richardson [10] demonstrated that a DL model implemented on the ARM® Cortex-M4 processor achieved superior classification performance compared to simpler algorithms, such as the linear discriminant analysis (LDA) classifier.

Thus, to effectively implement RNS in clinical settings, it is crucial to maintain high seizure detection performance while operating within the constraints of limited hardware resources. Furthermore, minimizing the number of implanted ECoG electrodes is essential, and the optimal placement of these electrodes must be systematically determined for each individual patient. This study aims to systematically reduce computational costs and the number of ECoG electrode channels while maintaining performance in patient-specific DL-based seizure detection. In this study, we used a 3D-CNN model for seizure detection. First, we trained a 3D-CNN on 3D ECoG data and analyzed the spatial and temporal characteristics of the learned first convolutional kernel. This analysis enabled us to systematically select the four channels that most contributed to seizure detection, determine the optimal number of kernels, and reduce the number of CNN parameters. We compared this systematic channel selection approach with manual channel selection. Finally, we evaluated the performance of a 2D-CNN model trained with the selected channels and kernel parameters, verifying whether real-time seizure detection could be achieved with reduced computational complexity.

## II. Materials and methods

### A. Clinical dataset

epilepsy patients, publicly available from IEEG.org [13][14]. This database contains annotations of seizure onset and offset made by clinical experts, making it well-suited for the development of seizure detection models. In this study, we defined the period from the onset to the offset of a seizure as the “ictal period,” the 30 minutes preceding the seizure as the “preictal period,” the 30 minutes following the seizure as the “postictal peripd,” and all other periods as the “interictal period” (Fig.1).

**Fig. 1.**
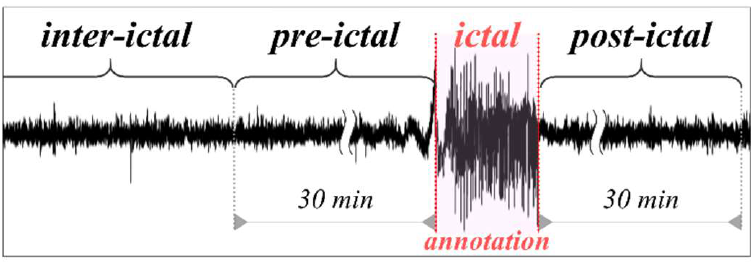
An example of continuous ECoG recordings and the four segments based on annotations by clinical experts.

We selected datasets from five patients. Here, the five patients are referred to as Patients A, B, C, D, and E. The study utilized ECoG data sampled at various frequencies (499.917Hz, 500Hz, 5000Hz) with recording durations spanning one to six days. To align with our 3D-CNN architecture (detailed in Section II-B), all ECoG signals were downsampled to a unified sampling rate of 256Hz. No preprocessing steps, including high-frequency noise filtration or low-frequency drift compensation, were implemented. For each patient, all channels of the intracranial grid electrodes (Fig. 2) were utilized, with electrode placement information preserved. Patient A used a 6×6 grid with 36 channels, patient B used a 4×6 grid with 24 channels, patient C used an 8×8 grid with 64 channels, patient D used an 8×3 grid with 24 channels, and patient E used an 8×8 grid with 64 channels. For patient A, ECoG signal data for electrode channels #1, 17, 18, 35, and 36 were missing, and these channels were filled with zero values.

**Fig. 2.**
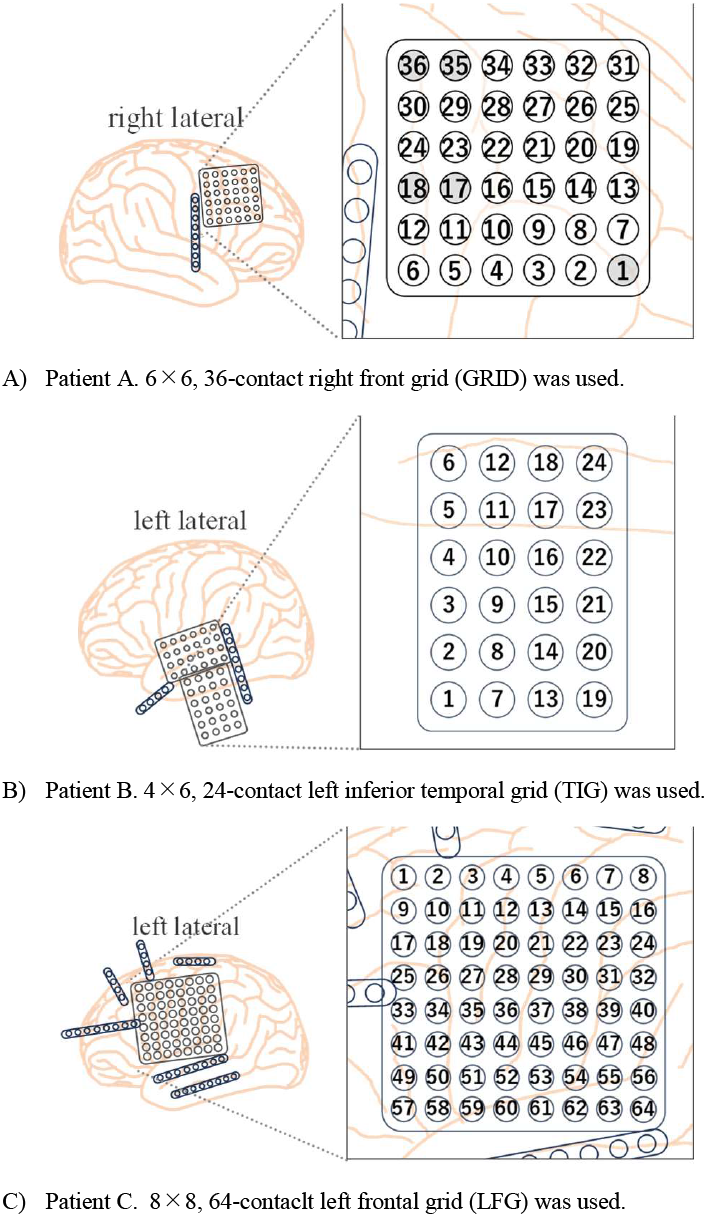
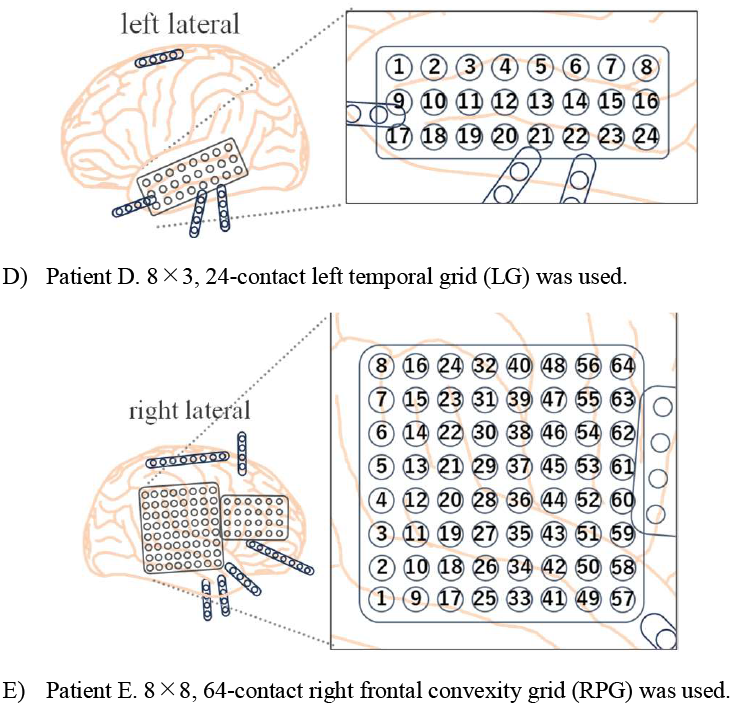
Intracranial electrode placements in five patients [15] and the grid electrode channel configurations used in this study.

### B. Neural network structure

In this study, we developed a 3D-CNN architecture based on the SeizureNet CNN [11] for patient-specific real-time seizure prediction. The detailed network structure of this model is presented in TABLE I. The model input is structured as a four-dimensional tensor. The first axis represents the vertical spatial dimension, corresponding to the vertical electrode channel arrangement in the ECoG data. The second axis represents the horizontal spatial dimension, reflecting the horizontal electrode channel layout. The third axis is the temporal dimension, indicating the temporal variation of the ECoG signals. The fourth axis denotes the number of channels, which is treated as a single channel here and processed as a grayscale image. The proposed 3D-CNN consists of five convolutional layers. In the initial 3D convolution stage, 20 parallel temporal convolution layers are used to extract both spatial and temporal patterns from the 3D-ECoG data (Fig. 3). In the time convolution layer, kernels with a size of (1×1×17) are arranged according to the grid electrode channel configuration. Each kernel extracts temporal features from a specific electrode channel. The output of this convolution layer is reshaped from 3D to 2D through dimensional transformation. After the initial 3D convolution, feature extraction is performed using four 2D convolution layers. Each convolution layer performs temporal convolution with a kernel size of (1×5). In all convolutions, the stride is set to 1, and no padding is applied. Batch normalization is applied to stabilize the output distribution of each layer, accelerating training and improving model accuracy. The ReLU activation function is used to introduce non-linearity while mitigating the vanishing gradient problem. Pooling layers are placed after each convolutional layer to reduce the spatial size of the data and aggregate features. Max pooling is used for downsampling in the time direction. The first and second pooling layers use a pool size of (1×4), reducing the time direction size by a factor of 4. The third pooling layer uses a pool size of (1×2), compressing the size by half. During training, dropout is introduced to prevent overfitting, improving the model’s generalization performance by randomly deactivating neurons. In the final 2D convolutional layer, the number of kernels is set to 2 with a kernel size of (1×1) to reduce the dimensions of the feature map for final classification. Then, a Flatten layer is used to convert the output of the convolution layers into a one-dimensional vector, preparing it for input into the fully connected layer. In the subsequent final layer, a fully connected layer and a softmax activation function are used to output the probabilities for the two classes: seizure and non-seizure states. For simplicity, a classification threshold of 0.5 is set during classification.

**TABLE I.**
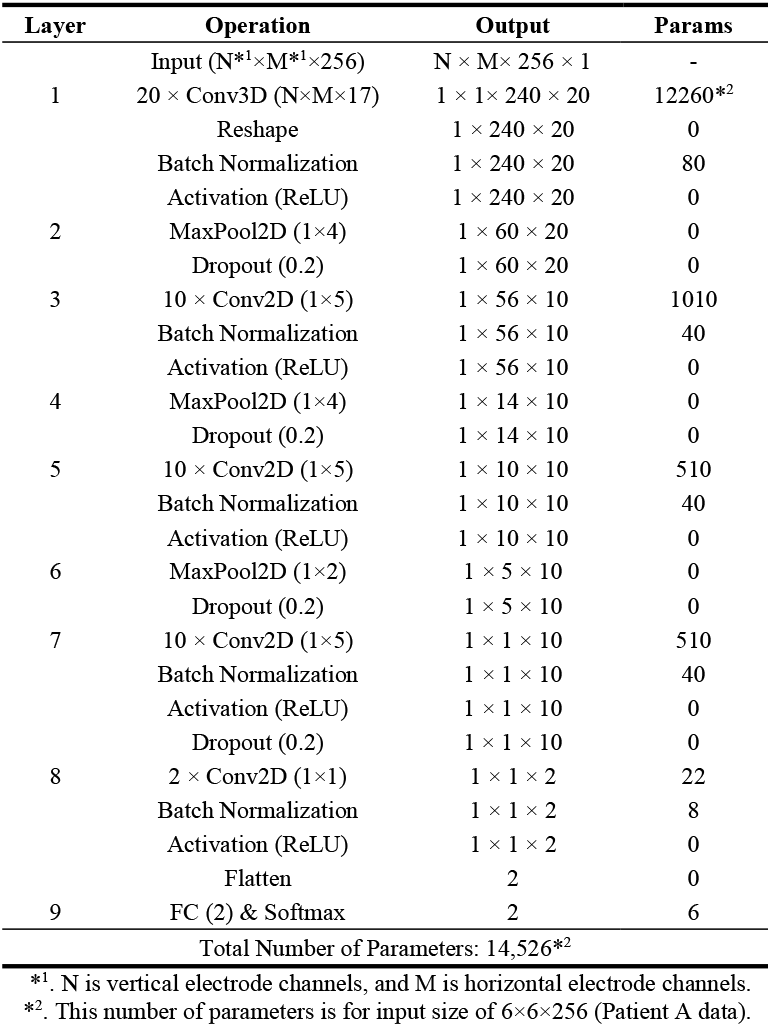
DETAILS OF 3D-CNN STRUCTURE.

| Layer | Operation | Output | Params |
| --- | --- | --- | --- |
| | Input ($N \times M \times 256$) | $N \times M \times 256 \times 1$ | - |
| 1 | $20 \times \text{Conv3D} (N \times M \times 17)$ | $1 \times 1 \times 240 \times 20$ | 12260*<sup>2</sup> |
| | Reshape | $1 \times 240 \times 20$ | 0 |
| | Batch Normalization | $1 \times 240 \times 20$ | 80 |
| | Activation (ReLU) | $1 \times 240 \times 20$ | 0 |
| 2 | MaxPool2D ($1 \times 4$) | $1 \times 60 \times 20$ | 0 |
| | Dropout (0.2) | $1 \times 60 \times 20$ | 0 |
| 3 | $10 \times \text{Conv2D} (1 \times 5)$ | $1 \times 56 \times 10$ | 1010 |
| | Batch Normalization | $1 \times 56 \times 10$ | 40 |
| | Activation (ReLU) | $1 \times 56 \times 10$ | 0 |
| 4 | MaxPool2D ($1 \times 4$) | $1 \times 14 \times 10$ | 0 |
| | Dropout (0.2) | $1 \times 14 \times 10$ | 0 |
| 5 | $10 \times \text{Conv2D} (1 \times 5)$ | $1 \times 10 \times 10$ | 510 |
| | Batch Normalization | $1 \times 10 \times 10$ | 40 |
| | Activation (ReLU) | $1 \times 10 \times 10$ | 0 |
| 6 | MaxPool2D ($1 \times 2$) | $1 \times 5 \times 10$ | 0 |
| | Dropout (0.2) | $1 \times 5 \times 10$ | 0 |
| 7 | $10 \times \text{Conv2D} (1 \times 5)$ | $1 \times 1 \times 10$ | 510 |
| | Batch Normalization | $1 \times 1 \times 10$ | 40 |
| | Activation (ReLU) | $1 \times 1 \times 10$ | 0 |
| | Dropout (0.2) | $1 \times 1 \times 10$ | 0 |
| 8 | $2 \times \text{Conv2D} (1 \times 1)$ | $1 \times 1 \times 2$ | 22 |
| | Batch Normalization | $1 \times 1 \times 2$ | 8 |
| | Activation (ReLU) | $1 \times 1 \times 2$ | 0 |
|  | Flatten | 2 | 0 |
| 9 | FC (2) & Softmax | 2 | 6 |
| Total Number of Parameters: 14,526*<sup>2</sup> |  |  |  |

\*<sup>1</sup>. N is vertical electrode channels, and M is horizontal electrode channels.
\*<sup>2</sup>. This number of parameters is for input size of $6 \times 6 \times 256$ (Patient A data).

**Fig. 3.**
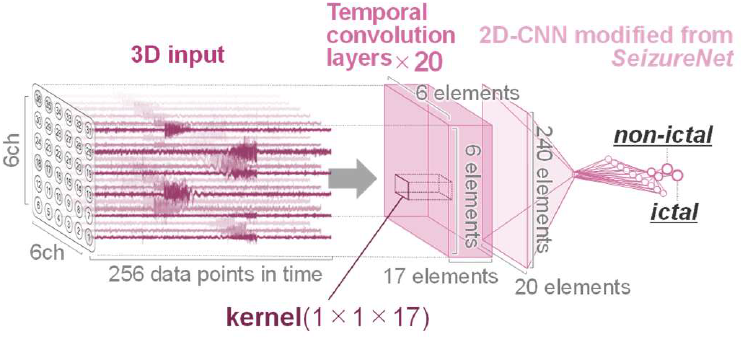
The 3D-CNN architecture (example: Patient A). The sizes of the 3D input and temporal convolution layers vary by patient.

To enable real-time seizure detection based on patient-specific ECoG signals, a 2D-CNN was adopted with reduced input size and kernel number for the first convolutional layer. The input to the 2D-CNN consists of one dimension along the time axis and another dimension corresponding to the four selected channels, as discussed later (Section II-G). Furthermore, the number of kernels in the first convolutional layer was optimized through the analysis of the temporal kernels in the 3D-CNN (Section II-H). The remaining structure follows the same configuration as the aforementioned 3D-CNN.

### C. Data preparation for CNN training and testing

In this study, specific criteria were established for selecting seizure events from the datasets of five patients. Seizure events were excluded from analysis if data from 15 minutes prior to the seizure onset were missing, if the intervals between seizures were too short, or if the recordings contained noise or artifacts. Ultimately, 14, 7, 19, 6, and 5 seizure events from patients A, B, C, D, and E, respectively, were included in the analysis.

For each seizure event, we extracted data segments from both the ictal and interictal periods (excluding the 30-minute pre-ictal and post-ictal periods). These segments were designed to match the input dimensions of the respective models: electrode channels × electrode channels × 256 samples for the 3d-CNN and 4 electrode channels × 256 samples for the 2d-CNN. A sliding window approach was employed, with a step size of 1 sample during seizures and 128 samples during interictal periods. However, for a specific seizure event in patient A, the limited interictal data necessitated a reduced step size of 6 samples for interictal period sampling.

For each seizure event, both ictal and interictal datasets were created. Afterward, the holdout method was used to split the sequential ECoG signal data into training and validation datasets. TABLE II presents the breakdown of data samples in the training and validation datasets for each patient obtained through this process.

**TABLE II.** BREAKDOWN OF DATA SAMPLES.

| Patient I.D. |  | inter-ictal | ictal | Total |
| --- | --- | --- | --- | --- |
| A | for train | 32,9186 | 65,381 | 39,4567 |
|  | for validation | 62,8833 | 45,106 | 67,3939 |
| B | for train | 12,2440 | 5,8306 | 18,0746 |
|  | for validation | 14,1726 | 3,3881 | 17,5607 |
| C | for train | 80,822 | 112,266 | 193,088 |
|  | for validation | 155,896 | 51,877 | 207,773 |
| D | for train | 115,331 | 84,842 | 200,173 |
|  | for validation | 62,802 | 48,731 | 111,533 |
| E | for train | 127,406 | 33,251 | 160,657 |
|  | for validation | 94,433 | 25,999 | 120,432 |

The test dataset for the trained 2d-CNN model comprised two data types.

Seizure Episode Data: 4-channel ECoG signals recorded 15 minutes pre-ictal period, throughout the entire ictal period, and 5 minutes post-ictal period.

Non-Seizure Resting State Data: 4-channel ECoG signals recorded during a continuous one-hour period within an interictal period exceeding nine hours.

The number of seizure episode data samples varied per patient based on the number of recorded seizures, while each patient contributed a single non-seizure resting state data sample.

### D. Training of 3D-CNN and 2D-CNN

The 3D-CNN model was trained using each patient’s dataset. Training was conducted over 1200 epochs with a batch size of 512. The Adam optimizer [16] was employed with a learning rate set at 0.001. Sparse categorical cross-entropy, suitable for multi-class classification problems, was used as the loss function. To mitigate the vanishing gradient issue, He initialization [17] was applied for model initialization. Additionally, to evaluate model stability, training was repeated 12 times using different random seeds, resulting in 12 distinct models per patient.

Due to the imbalance in the number of samples between seizure and non-seizure periods in the dataset (see TABLE II), it was necessary to address the class imbalance issue during the training of the 3D-CNN. In this study, higher weights were assigned to the seizure period data, which is the minority class, to ensure the model learned both classes effectively. The class weights were set proportional to the inverse of the sample size for each class, as defined in Equation (1). This approach allowed the model to focus on improving the classification accuracy for the seizure period data.

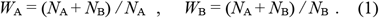

The 2D-CNN was trained using the same hyperparameters as the 3D-CNN. In contrast, only six different seed values were utilized for initialization. This resulted in the creation of six unique 2D-CNN models for each patient.

### E. Testing of post-trained 2D-CNN models

A sliding window with a size corresponding to the 2D-CNN input layer (4 electrode channels × 256 samples) was applied to both the seizure episode data and the non-seizure resting data. This window was shifted one sample at a time along the temporal axis, and the data within each window was fed into the 2D-CNN to estimate the presence or absence of seizures. However, no estimation was performed for the first 255 samples due to insufficient data size.

### F. Evaluation metrics

Four evaluation metrics were used to assess the inference performance of the trained 2D-CNN model.

The inference performance on seizure episode data was evaluated through the analysis of the area under the receiver operating characteristic curve (ROC-AUC) and the area under the precision-recall curve (PR-AUC). These metrics were calculated for each seizure event, resulting in a different number of ROC-AUC and PR-AUC values (*n* = number of seizure events × 6 seed values) for each patient.

In this study, a detection was defined as successful if the 2D-CNN model inferred an ictal state at least once during the ictal period. When detection was successful, detection delay was measured as the temporal discrepancy between the seizure onset time annotated by clinical experts and the moment when the estimated seizure state probability exceeded the threshold of 0.5. If detection was successful for all seizure event data, the number of detection delay calculations *n* was the same as the *n* for ROC-AUC and PR-AUC. However, if detection failed, detection delay was not calculated, resulting in a smaller *n*.

For the non-seizure resting state data, the performance evaluation metric used was the false positive rate (FPR), which was calculated as the proportion of time within a 1-hour interictal period where the estimated seizure state probability exceeded the threshold of 0.5. Since one non-seizure resting state data set was prepared for each patient, the number of FPR calculations *n* was *n=6* (1 data set × 6 seed values).

### G. Channel Selection

This section describes the manual channel selection method employed in our previous study [18] and the systematic channel selection method proposed in this study.

First, the manual channel selection method is described. For each patient, 8 channels were manually selected from the 24 to 64 channels of the intracranial grid electrodes based on the descriptions in the “iEEG Patient Report” database and the distinctive and important characteristics observed in the ECoG signal waveforms (e.g., relatively large amplitude or oscillatory activity before or during seizures). In previous studies [11][19], considering the limitations of the number of electrodes and the spatial range of implanted devices, 4 to 5 channels covering the seizure onset zone (SOZ) were chosen based on the definition of SOZ by clinical experts. However, since this study did not involve expert supervision, 8 channels were selected to ensure redundancy. For each patient, the selected set of 8 channels was as follows: for Patient A, channels #10, 11, 22, 23, 28, 29, 33, 34; for Patient B, channels #4, 5, 9, 10, 14, 15, 21, 22, 23; for Patient C, channels #28, 26, 36, 37, 42, 43, 50, 51; for Patient D, channels #1, 2, 10, 11, 12, 17, 18, 20; and for Patient E, channels #34, 35, 36, 37, 42, 46, 47, 51, 52. These channel set includes the 4 channels selected through training and testing of the convolutional autoencoder (CAE) model for seizure detection. The procedure for comprehensive channel selection using the CAE is outlined below.

1. A one-channel CAE was built for all channels, and the ROC-AUC was calculated. Channels that exceeded the threshold set for each seizure event were extracted.
2. All possible 3-channel combinations (nC3) were selected from the channel group chosen in 1.
3. A 3-channel CAE was then constructed for each of the selected 3-channel sets, and channels included in sets exceeding the threshold of 0.75 for all test seizure events were extracted.
4. From the channel group extracted in 3., all possible 4-channel combinations (mC4) were selected.
5. Finally, a 4-channel CAE was constructed for each of the selected 4-channel sets, and the set with the best performance was chosen.

Next, the systematic channel selection method proposed in this study will be described.

The results of the trained 3D-CNN model on the validation data for each patient are shown in TABLE III.

**TABLE III.** 3D-CNN TRAINING RESULTS FOR VALIDATION DATA.

| Patient I.D. | loss | accuracy |
| --- | --- | --- |
| *A* | 0.01308 - 0.08174 | 0.9969 - 0.9988 |
| *B* | 2.445 - 16.76 | 0.8090 - 0.8224 |
| *C* | 0.6086 - 3.552 | 0.9133 - 0.9561 |
| *D* | 0.3708 - 1.216 | 0.9397 - 0.9680 |
| *E* | 1.474 - 4.593 | 0.8540 - 0.8806 |

To select the electrode channels most contributing to seizure detection, the time convolution layer in the initial convolutional stage of the trained 3D-CNN model (Fig. 3) was used. The time convolution layer consists of (1×1×17) kernels arranged according to the grid electrode channel configuration. The absolute weights of each kernel were summed along the time axis and averaged across 20 parallel layers to create spatial heatmaps of the kernels for each of the 12 models per patient. Additionally, the kernel maps of the 12 models were averaged (Fig. 4). Based on this map, the four electrode channels with the highest values were selected as the most contributory to seizure detection.

**Fig. 4.**
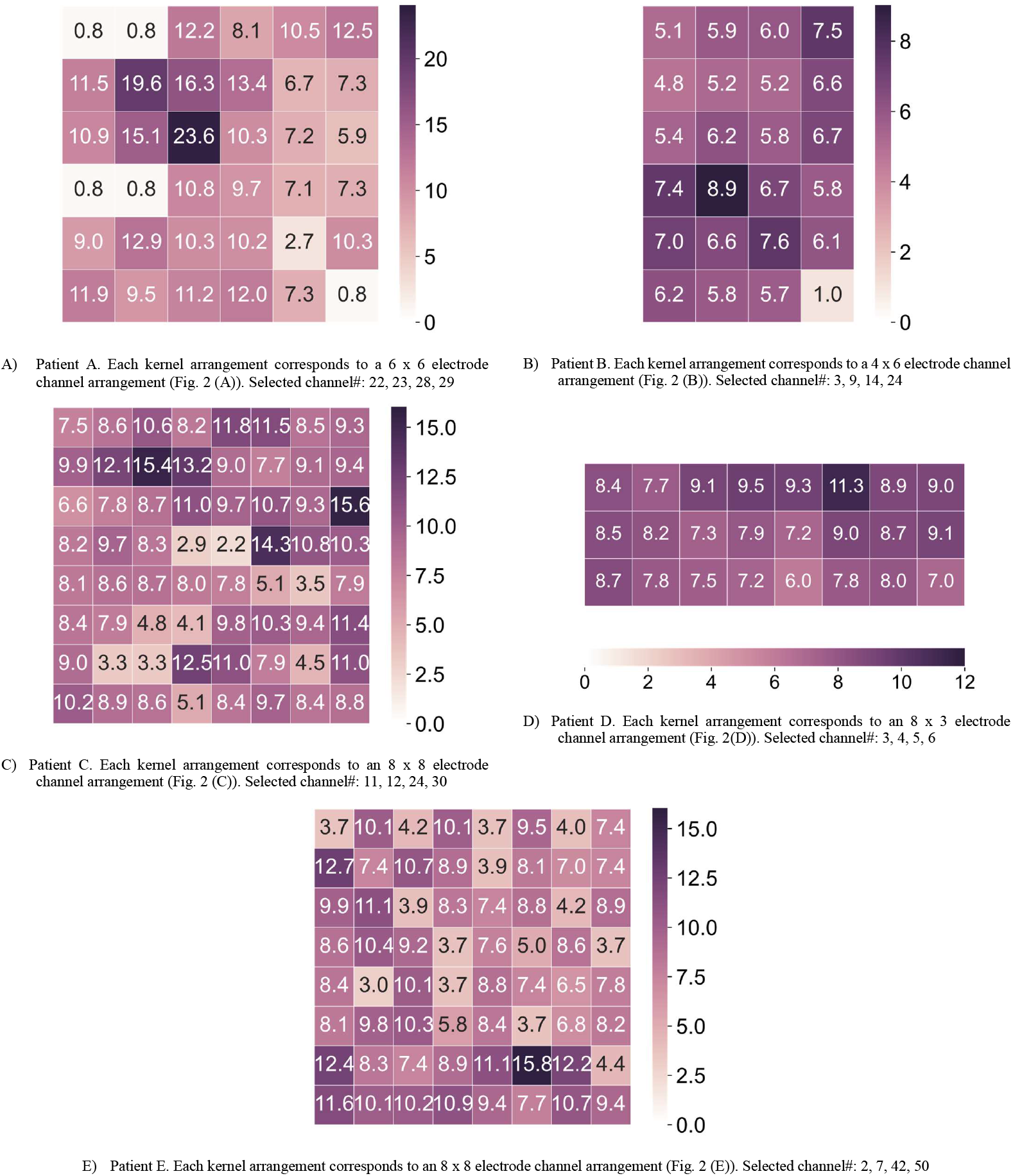
Spatial heatmaps of the kernel weights in the first convolutional layer of the trained 3D-CNN for five patients, averaged over 12 models. The range of the color bar differs for each patient.

### H. Temporal convolutional layers

The kernel weight values of the first convolutional stage for the selected four effective ECoG electrode channels were visualized as time waveforms (Fig. 5a). Not all of the 20 parallel layers exhibited temporal features, but several layers showed characteristic kernel weight values with temporal patterns. Therefore, the number of required parallel layers in the first convolutional stage was determined using the following procedure.

**Fig. 5.**
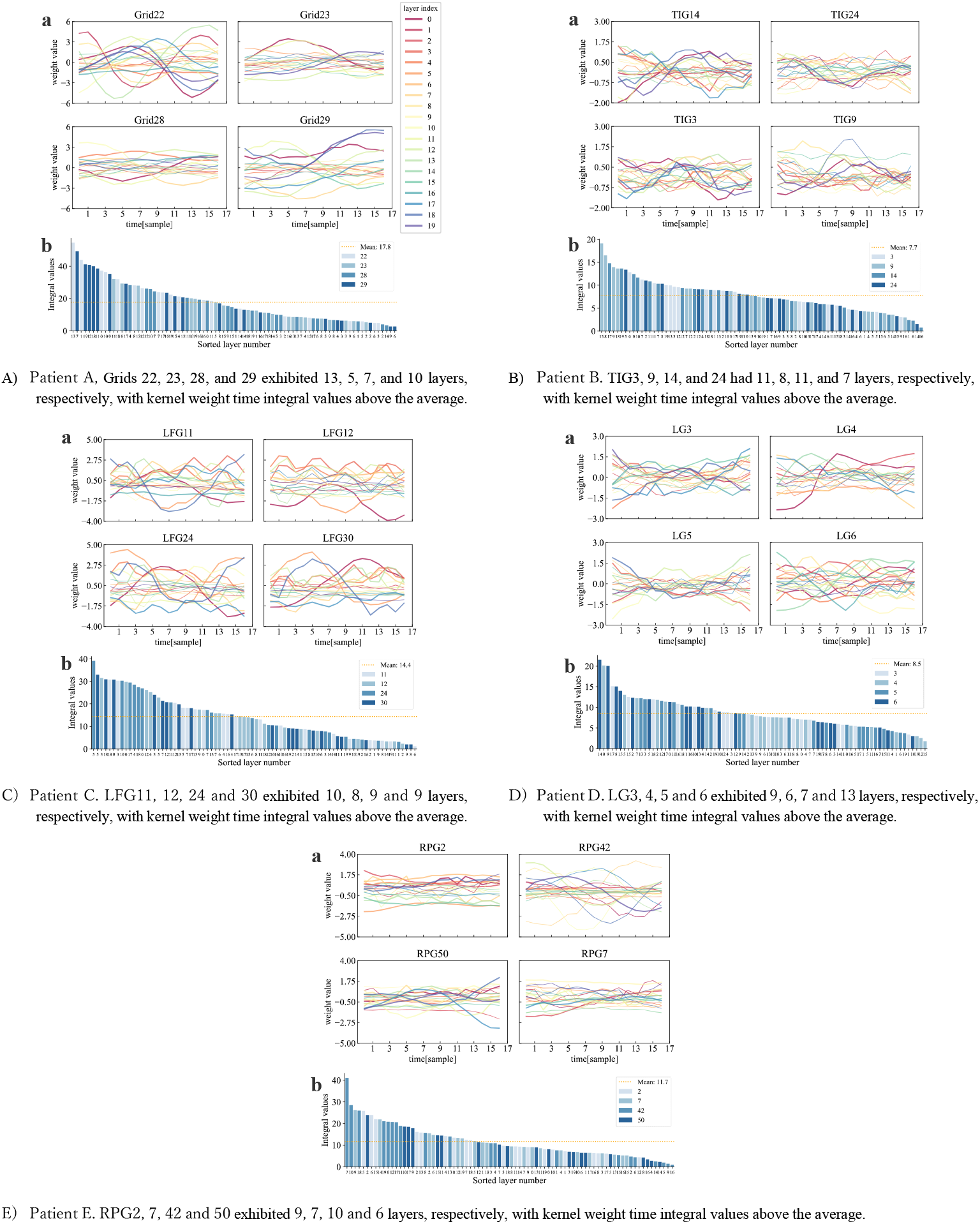
(a) Visualizes kernel weight values for each of the 20 parallel layers in the first convolutional stage of the trained 3d-CNN models across five patients. Layer indices 0-19 are color-coded. Layers with above-average absolute kernel weight time integrals are highlighted with thicker lines. (b) Shows the absolute kernel weights for all 80 layers sorted in descending order. Four channels are represented by different colors. The horizontal axis represents sorted layer indices, and the vertical axis displays time integral values of absolute kernel weights. The dashed line indicates the average value.

1. Calculate the time integral of the absolute kernel weights for all 80 layers (20 parallel layers × 4 channels).
2. Sort the calculated time integral values in descending order (Fig. 5b).
3. Count the number of layers with values greater than or equal to the average for each channel.
4. The number of layers with the most occurrences across the 4 channels is taken as the required number of parallel layers per channel.

The required number of parallel layers determined by this procedure was 13, 11, 10, 13, and 10 for patients A, B, C, D, and E, respectively.

## III. Results

### A. 2D-CNN with the reduced number of parallel layers

The 2D-CNN was trained using data from four systematically selected electrode channels, as described in Section II-G, with the number of parallel layers in the first convolutional stage optimized according to Section II-H. The training results of all 2D-CNN models (5 patients × 6 models) on the validation data are presented in TABLE IV.

**TABLE IV.** 2D-CNN TRAINING DATA FOR VALIDATION DATA.

| Patient I.D. | loss | Accuracy |
| --- | --- | --- |
| A | 0.0166 - 0.0483 | 0.9949 - 0.9985 |
| B | 2.371 - 34.744 | 0.8079 - 0.8092 |
| C | 0.2657 - 0.6428 | 0.9682 - 0.9778 |
| D | 0.0708 - 0.6029 | 0.8543 - 0.9868 |
| E | 0.3149 - 1.333 | 0.8622 - 0.9265 |

We compared the inference performance of the 2D-CNN model proposed in our previous study [18], which used data from 8 manually selected electrode channels and featured 20 parallel layers in the first convolutional stage, with that of the proposed downsized 2D-CNN model after training.

Fig. 6a-d compare the inference performance of the previous 2D-CNN model (blue plot) and the proposed downsized 2D-CNN model (orange plot) across five patients, using the four evaluation metrics described in Section II-F.

**Fig. 6.**
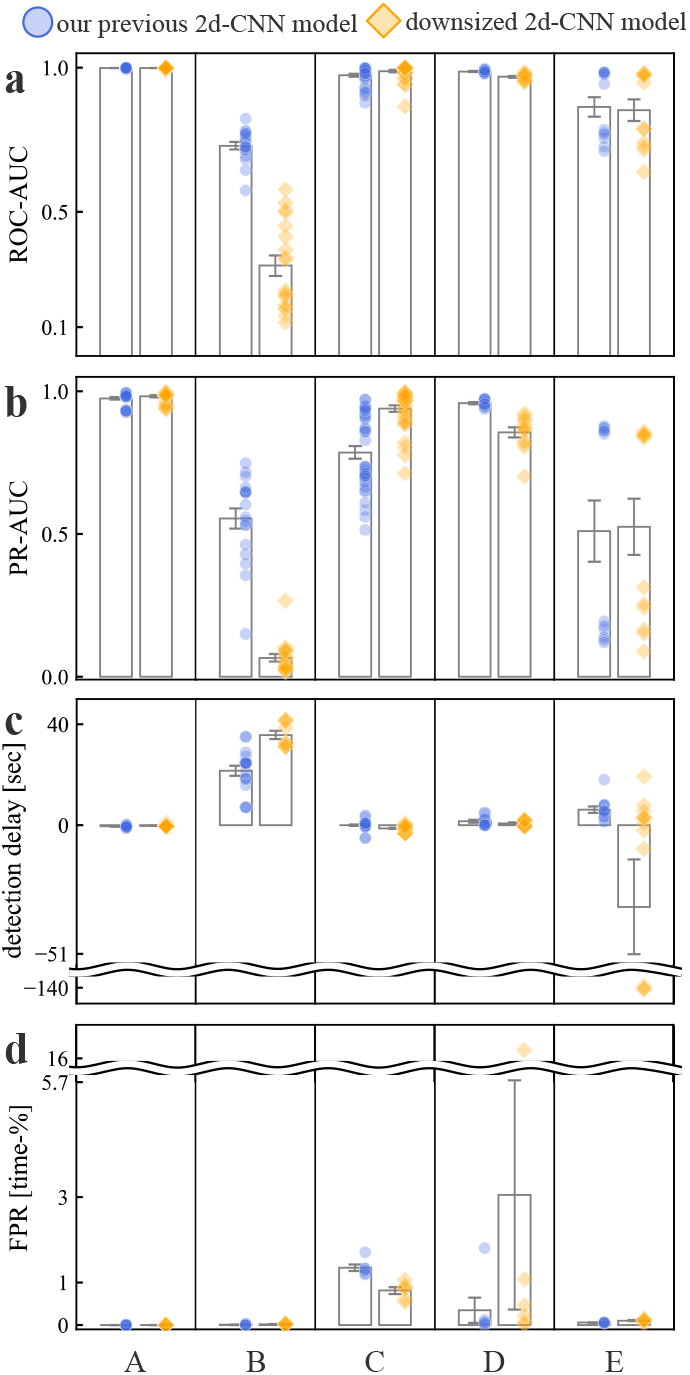
(a) ROC-AUC, (b) PR-AUC, (c) Detection delay, and (d) FPR. The bar graphs represent the mean, and the error bars indicate the standard error. The sample sizes for ROC-AUC and PR-AUC were n=30, 18, 36, 12, and 12 for patients A, B, C, D, and E, respectively. The sample size for detection delay is as mentioned above; however, for patient B, detection delay occurred only in the downsized model (orange plot), so n=9. The sample size for FPR was n=6 for all patients.

For patient A, both the previous 2D-CNN model and the downsized 2D-CNN model demonstrated superior performance compared to other patients. These models achieved high scores across the four evaluation metrics, with no significant differences observed between them. Additionally, there was minimal performance variation among the six models trained under different initialization conditions.

The 2D-CNN model constructed using data from patient B exhibited lower seizure detection accuracy compared to other patients. The model’s performance was susceptible to the type of seizure and the random seed used during initialization, resulting in a lack of stability. The model for patient B showed moderate accuracy during the training phase (TABLE IV), but its performance during the test phase was unsatisfactory. This was because, while the model accurately predicted non-seizure resting state data, it failed to correctly detect seizure episodes. Consequently, the ROC and PR AUC scores were significantly low, detection latency was substantially high, and the false positive rate was close to 0%. This suggests that the training method employed in this study may not be universally applicable to all patients.

Patients C and D showed favorable results, following patient A, but there was significant variation in the FPR among the six models trained with different seed values. For patient C, the downsized 2D-CNN model outperformed the previous 2D-CNN model across all evaluation metrics.

For patient E, the inference performance varied significantly depending on the type of seizure episode. Both the previous and downsized models exhibited notably low PR-AUC for certain seizure events, failing to make accurate predictions. Therefore, it seems challenging to improve the performance of the patient-specific 2D-CNN model using the same seizure event data.

The negative detection delay in patients A, C, and E suggests that there may be signals with characteristics similar to those of the ictal period during the pre-ictal period, indicating the possibility of estimating a ictal onset time that differs from the judgment of medical professionals. Alternatively, it may suggest the possibility of seizure prediction. The performance evaluation results revealed that downsizing the 2D-CNN model did not significantly affect the model’s performance, except for patient B.

## I. Disucussion

The method proposed in this study was able to reduce the number of ECoG electrode channels and the number of parallel layers in the first convolutional stage of the 2d-CNN for immediate seizure detection, while maintaining validation accuracy in at least some patients. TABLE V presents a comparison between the previous 2d-CNN model and the downsized 2d-CNN model, showing the number of electrode channels and the number of parameters in the 2d-CNN.

**TABLE V.** Changes in the number of channels and NN parameters.

| Patient<br>I.D. code | Electrode channels |  | 2d-CNN parameters |  |
| --- | --- | --- | --- | --- |
|  | Previous<br>models | Downsized<br>models | Previous<br>models | Downsized<br>models |
| A | 36 | 4 | 3,646 | 2,785 |
| B | 24 |  |  | 2,539 |
| C | 64 |  |  | 2,416 |
| D | 24 |  |  | 2,785 |
| E | 64 |  |  | 2,416 |

As shown in TABLE V, the input size of the CNN was reduced by 87.5% for patient A, 83.33% for patients B and D, and 93.75% for patients C and E. Additionally, the number of CNN parameters decreased by 23.61% for patients A and D, 30.36% for patient B, and 33.74% for patients C and E. Furthermore, in most cases excluding patient B, the inference accuracy was maintained.

In this study, we implemented a 2d-CNN-based seizure detector using 4 ECoG electrode channels on Google’s Edge TPU (Tensor Processing Unit) [20], and validated its real-time output by using the LED lighting behavior as a substitute for electrical signals to the head-implanted stimulation device. As a result, seizures were detected in real time in the annotated regions by clinical experts (see Fig. 1), and the LED lighting was confirmed. This demonstrated that the entire system, including the input of ECoG data, anomaly detection, and the output of electrical signals to the stimulation device, can process data in real time.

These findings demonstrate the potential of the proposed method for effectively minimizing hardware resource demands when deploying CNN-based seizure detection algorithms on resource-constrained devices, such as RNS.

## Data Availability

All data produced in the present study are available upon reasonable request to the authors.

https://bitbucket.org/ieeg/ieeg/wiki/Downloads.md

https://www.ieeg.org/

## Acknowledgments

This research was supported in part by Grant-in-Aid for Scientific Research from MEXT, Japan (22K12781 to YH).

